# Evaluation of 1^st^ WHO Anti-Malaria Reference Reagent for Competition ELISA Harmonisation and Development of ADAMSEL Analytical Platform

**DOI:** 10.1101/2023.10.16.23296996

**Authors:** Bhagwati Khatri, Peggy Riese, Hanna Shkarlet, Daniella Mortier, Helen McShane, Paul Bowyer, Edmond Remarque

## Abstract

This study focuses on harmonising the competition ELISA (cELISA) assay for *Plasmodium falciparum* (*P. falciparum*), using the 1^st^ WHO reference reagent for anti-malaria (*P. falciparum*) human reference serum (10/198). Antibody-mediated immune responses, against the Apical Membrane Antigen 1 (AMA1), play a significant role in protection against malaria. However, the sequence diversity of AMA1 and cross-reactivity among variants pose challenges in assessing antibody responses. To address this, the cELISA assay was selected to examine cross-reactive antibody responses against different variants.

The harmonisation process for cELISA was performed in three laboratories. The 10/198 served as an internal standard for the calculation of IgG concentrations in the cELISA using ADAMSEL software. Additionally, a novel semi-automated analytical tool was developed in the R-statistics environment. This tool is freely available for download and streamlines result in generation while minimising human error.

This study demonstrated the effectiveness of the 1^st^ WHO reference reagent as a standard for ELISA. Additionally, the ADAMSEL software and R-platform tool provide a user-friendly and accessible tool for the analysis of cELISA data. Its automation capabilities improve efficiency and ensure global accessibility at no cost, benefitting laboratories with limited resources.

**Importance:** This study focuses on the 1^st^ WHO reference reagent (10/198) in harmonizing the competition ELISA (cELISA) assay for P. falciparum. In addition, the introduction of ADAMSEL software for cELISA data analysis, coupled with a novel R-platform tool, available for free, simplifies analytical processes and contributes to global accessibility. This study demonstrates the 1^st^ WHO reference reagent’s efficacy as a competition ELISA standard.

## Introduction

Harmonisation of assays is essential in the field of preclinical vaccine development and the evaluation of vaccine candidates in clinical trials. By harmonising assays, researchers can ensure the comparability, reproducibility and robustness of results obtained from different laboratories, leading to improved patient outcomes and enhanced safety. Hence, harmonisation plays a crucial role in informing the outcome of vaccine trials.

One particular area where assay harmonisation is crucial is in the study of *Plasmodium falciparum* (*P. falciparum*), the most severe form of malaria which is responsible for 97% of malaria-related deaths globally [1]. While there is a malaria vaccine targeting the pre-erythrocytic stage that exhibits moderate efficacy [2], antibody-mediated immune responses play a vital role in protection against malaria [3-6]. Apical Membrane Antigen 1 (AMA1) is the ectodomain, micronemal protein of the *P. falciparum* [7,8] that is essential during the invasion of host cells [7,9-11]. The specificity of antibody responses to the polymorphic malaria antigen AMA1 can be broadened by immunisation with a limited number of antigenic variants [7,8,12,13]. However, AMA1 is polymorphic and exhibits sequence diversity and 50% of the IgG response after vaccination cross-reacts with AMA1 variants [14]. Therefore, to assess the cross-reactive antibody responses against heterologous variants, competition ELISA (cELISA) is useful as an *in vitro* assay to evaluate the breadth of the immune response and provide information on vaccine efficacy.

In this study, the cELISA assay was chosen for harmonisation due to its adaptability to diseases with polymorphic antigens. Additionally, several considerations were made before undertaking cELISA harmonisation across different laboratories. Participation in the harmonisation process was based on laboratories’ experience in conducting relevant immunological assays and considering factors such as available resources and shipment logistics. The study was funded by the FP7-INFRA-2012 Grant Agreement No. 312661, EURIPRED (European Research Infrastructures for Poverty-Related Diseases) [15]. One of the key activities of this project was to provide free access to high-quality reference reagents, scientific services, training courses, assay harmonisation and standardisation.

To ensure data normalisation, the 1^st^ WHO reference reagent for anti-malaria (*P. falciparum*) human serum (10/198) was used as an internal standard in the cELISA assay. This study introduces the ADAMSEL software package, and a semi-automated analytical tool in the R-statistics environment to calculate slopes and IC_50_ values emanating from the cELISA results.

Overall, this study focused on harmonising the cELISA assay for malaria but also demonstrated the applicability of the 1^st^ WHO reference reagent for anti-malaria (*P. falciparum*) human serum as a standard for AMA1 IgG. Additionally, this demonstrates the utility of the ADAMSEL software package. The downstream processing of cELISA results is further streamlined in the R statistics software environment [16], thereby reducing the time required for generating results and minimising the potential for human error. Importantly, the ADAMSEL and the downstream processing in R were developed with the intention of making it accessible to researchers globally at no cost, ensuring that laboratories with limited resources can benefit from its functionalities.

## Materials and Methods

### 10/198 serum and plasma samples

#### 10/198 serum

The 1^st^ WHO Reference Reagent for Anti-malaria (*P. falciparum*) human serum was developed in 2014 [17] [NIBSC code: 10/198, <u>Anti-malaria (Plasmodium falciparum) human serum(1st WHO Reference Reagent) (nibsc.org)</u>]. The 10/198 serum is a pooled plasma from 149 adult Kenyan individuals with a history of malaria [17]. Samples were screened for antibodies against five *P. falciparum*, MSP-1-19 (K1), MSP-1-42 (3D7), MSP-2 (3D7), MSP-3 (K1), AMA-1 (3D7, FC27, FP3), 77% positive for at least one antigen. Each ampoule contains a freeze-dried residue of diluted human plasma containing antibodies to *P. falciparum*. The freeze-dried ampoule of 10/198 serum was reconstituted in 1 ml of sterile water and used in subsequent assays or stored at -20^0^C. The plasma samples 28 and 32 were from malaria-reactive plasma packs stored at NIBSC. These samples were obtained from individuals from the United Kingdom and supplied by the National Health and Services, blood and Transplant unit (www.nhsbt.nhs.uk).

### cELISA

The cELISA protocol utilised in this study was primarily based on the publication by Kusi, et al. [13], with only one modification regarding the conjugate used. Instead of using Alkaline Phosphatase conjugate, as described in the published method by Kusi, et al.[13], this study used a Horseradish-Peroxidase-Conjugate (HRP). Four AMA1 antigens, namely HB3, 3D7, and DiCo 1 and 3, were selected for the cELISA assay. The choice of these specific AMA1 antigens against the 10/198 serum was based on their specificity and availability, as depicted in Supplementary Figure 1.

#### Step 1 (Fig. 1): Serum/patients’ plasma dilution determination against AMA1 antigens

**Figure 1:**
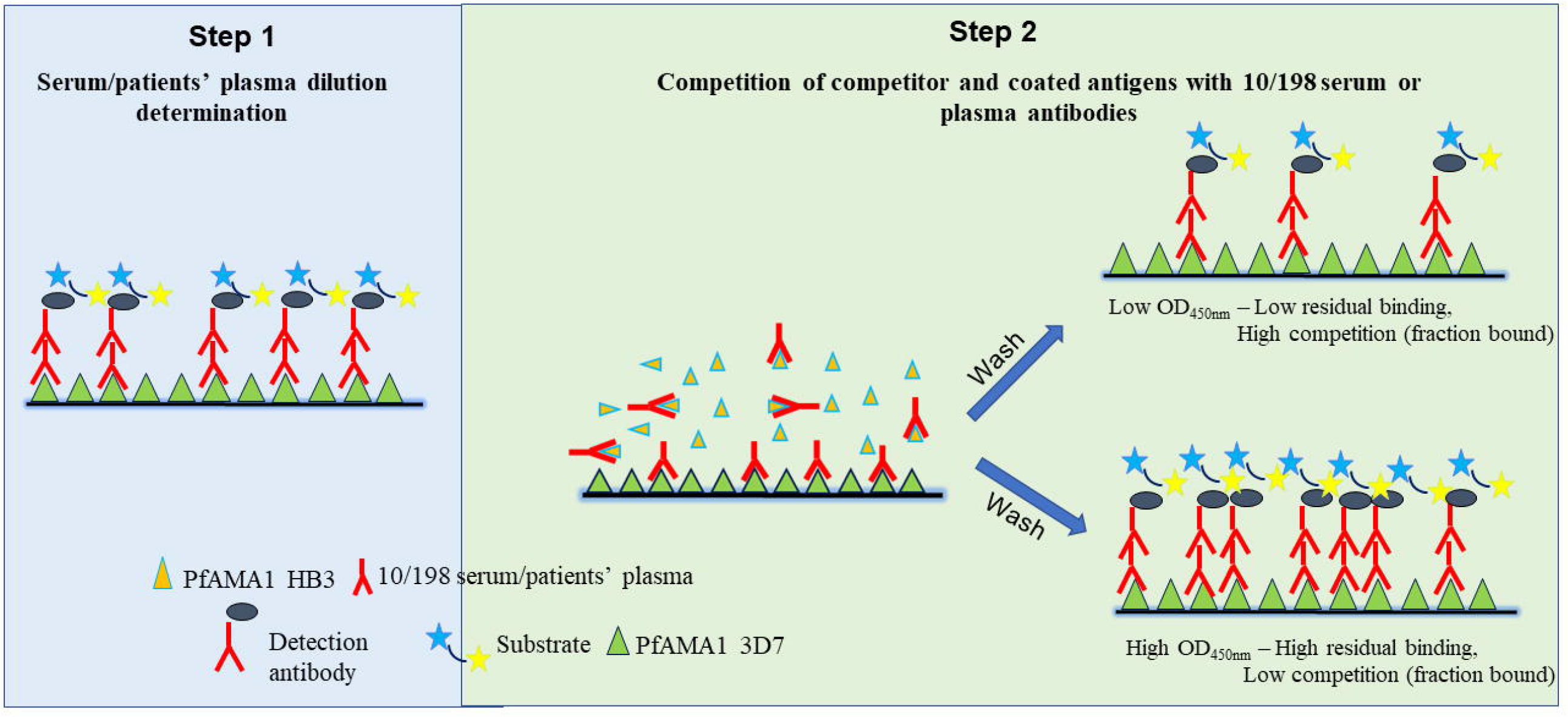
A schematic diagram illustrates the sequential steps involved in conducting a cELISA using the example of 3D7 as the coated antigen and HB3 as the competitor antigen. Briefly, step 1 involves choosing the optimal serum dilution of serum/patients’ plasma against each coated antigen (3D7). An optimal dilution of serum or plasma from step 1 is used to compete with the competitor antigen, HB3 antigen, in this case, for binding to the 3D7 antigen. Subsequent washing eliminates any unbound components, allowing for the detection of competition between the coated and competitor antigens. The detection antibody is then quantified using a spectrophotometer. The intensity of the signal is inversely proportional to the amount of competition that occurred between the coated antigen (3D7) and the competitor antigen (HB3). By comparing the signal intensity to a standard curve or known controls, the level of competition and the presence of specific antibodies can be determined.

An initial titration was performed to determine the optimal dilution of 10/198 serum or plasma required for the competition assay with each of the four antigens as coating antigens. Briefly, 96-well flat-bottom Microlon titre plates (Nunc MaxiSorp™ flat-bottom) were coated with 100 μL/well of 2 μg/ml 3D7, HB3 and DiCo 1 or 3 AMA1 ectodomain in coating buffer (50 mM Carbonate buffer, pH 9.6). The plates were then incubated at 4°C overnight.

Plates were washed six times with PBS + 0.05% Tween 20 (PBS-WB) using an automated plate washer. To prevent non-specific binding, the plates were blocked with 200 μL/well of 3% BSA in PBS-WB for at least 1.5 hrs. Plates were washed six times with PBS-WB. The 10/198 serum was titrated three-fold, starting from a 1:1250 dilution, while the plasma was titrated two-fold, starting from a 1:200 dilution and incubated for two hours at room temperature. Plates were washed six times with PBS-WB and subsequently incubated for 1 hour at room temperature with Horseradish Peroxidase (HRP)-Conjugated rabbit anti-human IgG (Dako, Denmark) diluted 5000 times in 0.5% BSA in PBS-WB. Plates were washed six times with PBS-WB and 1-Step™ Ultra TMB-Blotting substrate (Thermo Scientific, UK) was added, the colour reaction was stopped after 30 minutes by the addition of 2 M sulfuric acid and optical density (OD) values were read at 450 nm in an ELISA plate reader.

The 10/198 serum, titrated three-fold from a 1:1250 starting dilution, was used as a standard calibrator on all plates where patients’ plasma and 10/198 serum were used. All samples and standards were diluted with 0.5% BSA in PBS-WB and added in duplicate (100 μL/well).

As shown in Figure 2, two steps were employed for determining the dilution of the 10/198 serum and explained in more detail under the results section. Briefly, In Step 1a, serum dilution was determined based on achieving OD_450nm_ value of 1.5. Subsequently, each laboratory used their determined serum dilution in Step 2a. For Step 1b, a fixed dilution of the 10/198 serum was chosen and consistently used for Step 2b across all laboratories. For patients’ plasma samples, a fixed set of dilutions was employed during Step 2 of the cELISA assay.

**Figure 2:**
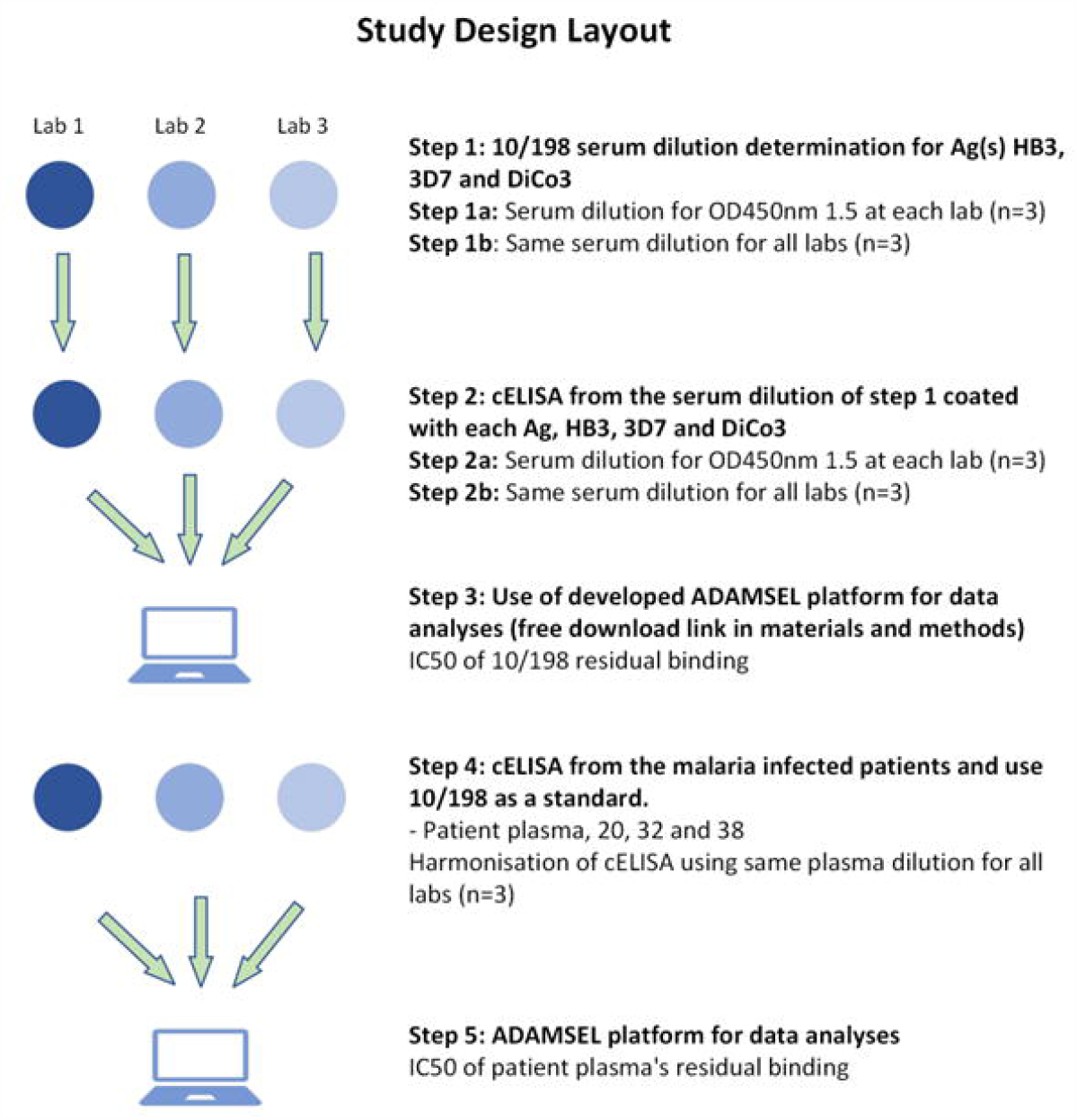
A schematic diagram of the cELISA harmonisation study design: Three laboratories performed cELISA using the same protocol in their respective laboratories. The 10/198 serum and antigens, HB3, 3D7 and DiCo3, were shared and used across the laboratories. The study design was divided into two stages. Stage 1 contained Steps 1 2 &3, with steps 1 and 2 were performed three times on separate occasions, triplicate experiments (n=3). The stage 2 contained Steps 4 and 5, Step 4 was performed by two laboratories 2 and 3 with duplicate and triplicate experiments, respectively Steps 3 and 5 were for data analyses using ADAMSEL software and R scripts.

### Step 2 (Fig. 1): Competition of competitor and coated antigens with 10/198 serum or plasma antibody

The assay involved the co-incubation of different allelic forms of *Pf*AMA1 (HB3, 3D7, DiCo 1 or 3) with the serum dilutions obtained from the Step 1a and Step 1b. Each laboratory performed three independent experiments for each *Pf*AMA1 allele. As a general principle, for the step 2, the plates were coated entirely with one of the variants of *Pf*AMA1 alleles, such that there was a competition between the added (competitor) antigens and 10/198 serum antibodies or patients’ plasma (dilution value was obtained from Step 1) for the binding to the coated antigens. For the Step 2 process, briefly, 96-well flat-bottom Microlon titre plates (Nunc MaxiSorp™ flat-bottom) were coated with 100 μL/well of 2 μg/ml of either of the four *Pf*AMA1 antigens in coating buffer (50 mM Carbonate buffer, pH 9.6). The plates were incubated at 4°C overnight and washed six times with PBS-WB using an automated plate washer. The plates were blocked with 200 μL/well of 3% BSA in PBS-WB for at least 1.5 hrs and washed six times with PBS-WB. The competitor antigens were serially titrated three-fold with 0.5% BSA in PBS-WB in the plates from 30 – 0.005 μg/mL, with seven duplicate wells for each concentration. The eighth well was left without competitor antigen. The 10/198 serum or patients’ plasma (at a two-fold higher desired dilution) was added to all 8 wells with or without competitor antigens. The two columns were used for the standard dilutions and 10/198 was used as a standard for all cELISA. For the standard dilutions, the 10/198 at 1 in 1250 dilution was added to the first 2 wells of the column and subsequently -three-fold serial dilution was performed up to 7 duplicate wells, the 8^th^ well was blank containing 0.5% BSA in PBS-WB. After two hours of incubation, the plates were developed as described in Step 1.

### ADAMSEL software and R scripts for analyses

To manage the substantial volume of data generated from the cELISA harmonisation study, the ADAMSEL software was developed. ADAMSEL is available for free download from the https://www.bprc.nl/sites/default/files/pubs/Adamsel-Suite.zip.

ADAMSEL was employed to calculate concentrations from the OD values using a four-parameter logistic curve fit on the 10/198 standard included on every plate, data were then exported and read into R to calculate the IC_50_ values for each coat-competitor combination. As a first step, the fraction of IgG remaining bound at various competitor concentrations was calculated by dividing the IgG concentration with competitor by the IgG concentration without competitor yielding values ranging between 0 and 1 (using the R scripts), followed by a least-squares regression utilising a two-parameter logistic function to estimate the IC_50_ and slope values. The R scripts and example data can be downloaded from : https://www.bprc.nl/sites/default/files/pubs/Data-and-Scripts.zip

The percent antibody depletion for any given competitor antigen is obtained by calculating the ratio between 100% (representing binding in the absence of competitor antigen) and the residual binding observed at the competitor concentration. The formula for the fraction remaining bound is as follows:

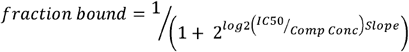

Where *fraction bound* is the remaining fraction at each competitor concentration, *IC*_*50*_ is the competitor concentration where 50% remains bound, *Comp Conc* is the competitor concentration and *Slope* is the steepness of the curve, with low values indicating steep curves. As the *Slope* parameter is somewhat difficult to interpret, it can also be expressed as the fold-increase in soluble antigen concentration required to change the fraction remaining bound from 90 to 50% or from 50 to 10%. This can be calculated by the following formula: 2^log2(1 /0.1 – 1) / -Slope^. In the event the estimated IC_50_ value was higher than twice the highest competitor concentration (viz. 30 μg/mL), it was arbitrarily set to 60 μg/mL to avoid extrapolation beyond the actual measurement range.

## Results

### cELISA principle

As depicted in Figure 1, the cELISA operates on the principle of assessing the ability of soluble competitor antigen(s) to compete with an immobilised coating antigen for binding to the target antibodies. The cELISA assay involves a two-step process. The initial step, Step 1 entails determining the optimal dilution of the serum or plasma sample against the coating antigen (i.e., an OD value of approximately 1.5). Step 2, involves incubation of the pre-determined sample dilution from Step 1 with a dilution series of the competitor antigens, either homologous or heterologous in the well containing the coating antigen. In Figure 1, for instance, the selected dilution of the 10/198 serum sample obtained from Step 1 is incubated with the competitor antigen HB3 in a well coated with the 3D7 antigen. By following this approach, the cELISA assay measures the competitive binding of the serum antibodies between the coated antigens and the soluble competitor antigens. The resulting intensity of the signal readout determines the degree of competition. For instance, if the 10/198 (Fig.1) yields a high OD_450nm_ signal, this suggests that serum antibody exhibits low competition towards the heterologous HB3 antigen with the coating antigen 3D7. Conversely, if a low OD_450nm_ signal is obtained, this indicates high competition of the serum antibody with the HB3 antigen suggestive of a high degree of cross-reactivity.

### Study Design

The study design for the harmonisation of the cELISA assay, as shown in Figure 2, involved three laboratories within the European continent. The harmonisation process was carried out in two stages. In the first stage, a serum dilution of 10/198 was employed as both a standard and a sample to establish the cELISA working protocol. This stage included familiarising the laboratories with the assay process, optimising the protocol, and conducting three independent cELISA assays at each of the three laboratories using the 10/198 serum and standard. The second stage of the harmonisation process aimed to validate the use of the 10/198 serum as a standard for the convalescent plasma sample obtained from patients who had previously contracted and successfully resolved malaria infections. These convalescent plasma samples were used as test samples in the cELISA assay to assess their reactivity and enabled a comparative analysis of the results obtained across the three laboratories. The data obtained from these assays were subsequently analysed using the ADAMSEL software and R.

By conducting the harmonisation study in these two stages, involving multiple laboratories and using both standard serum (10/198) and plasma samples, the aim was to ensure the robustness and reliability of the cELISA assay and validate its performance across different settings and sample types.

### cELISA harmonisation using 10/198 as a standard and serum

For the cELISA harmonisation, 10/198 was used as a standard and test serum. As shown in Fig 2, the cELISA steps 1 and 2 were subdivided into two further steps as Step 1a and 1b and Steps 2a and 2b.

#### Step 1

In Step 1a (Table 1a), a series of dilutions were prepared for the 10/198 serum, which were then incubated in plates coated with each antigen (HB3, 3D7, and DiCo3) at a concentration of 2 μg/mL. Each laboratory selected the 10/198 dilution based on the absorbance value of OD_450nm_ 1.5. For example, from Table 1a, in Lab 1, the 10/198 serum was subjected to a three-fold serial dilution and then incubated in a plate coated with HB3 antigen. The resulting OD_450nm_ values were plotted on a four-parameter logistic fit. From this analysis, the dilution of the 10/198 serum corresponding to an OD_450nm_ value of 1.5 was determined. In Lab 1, the 10/198 serum was found to be 1 in 17,000 times dilution. Similarly, Lab 2 achieved 1 in 38,000 times dilution, while Lab 3 had a dilution of 1 in 51,000 (Supplementary Figure 2).

**Table 1:**
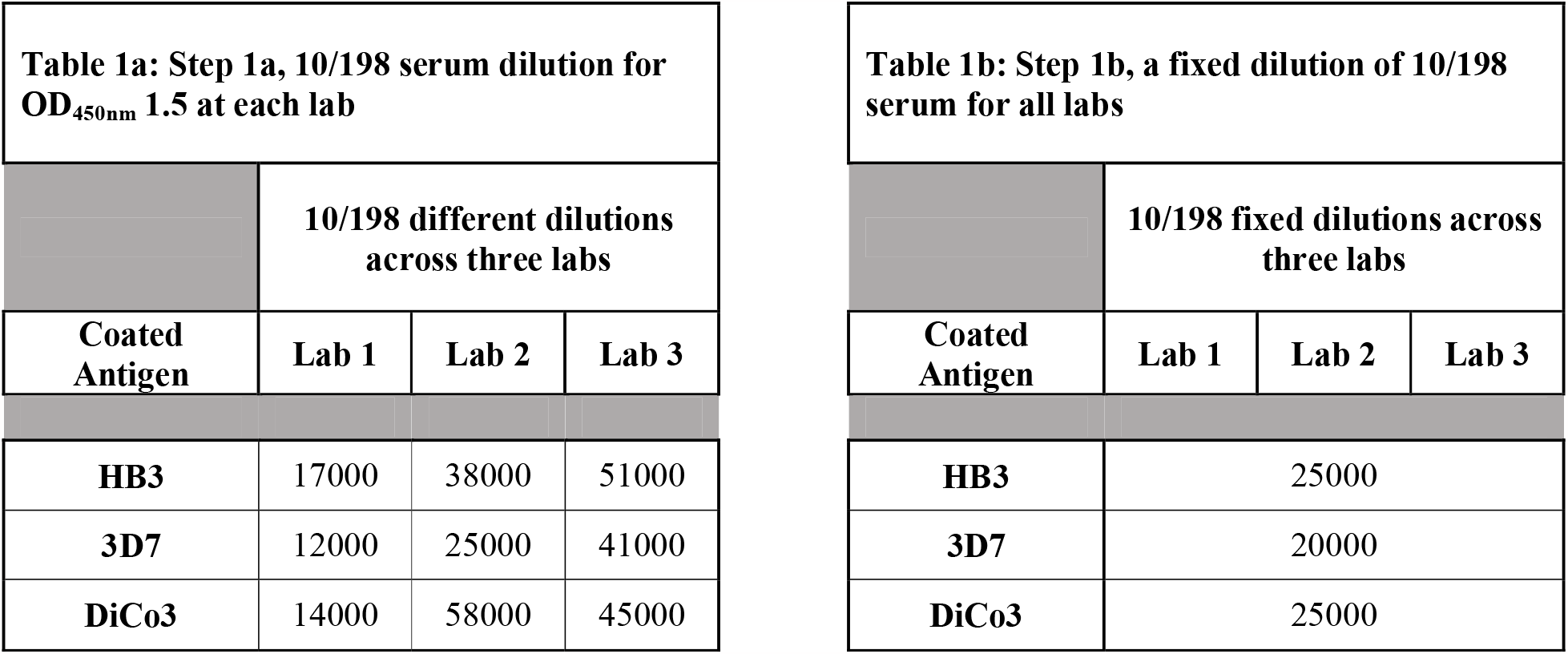
The chosen dilutions of 10/198 across three laboratories. Table 1a represents Step 1a in Fig. 2 and displays the different dilutions of 10/198 serum for each antigen obtained from the three laboratories. Notably, the dilutions of 10/198 serum for HB3 antigen coated plates vary, with dilutions of 1 in 17000, 1 in 38000, and 1 in 51000 for laboratories 1, 2, and 3, respectively. Table 1b, which represents Step 1b in Fig. 2, displays the identical dilution of 10/198 serum for each antigen derived from the linear portion of the standard curves from all three laboratories.

In Step 1b (Table 1b), a predetermined and standardised dilution of the 10/198 serum was selected and implemented across all three laboratories. The choice of this fixed dilution was based on the linear portion of the four-parameter logistic fit, ensuring consistency in the selection process. By examining the data from the four-parameter logistic fit, the dilution of the 10/198 serum that fell within the linear range was identified for each laboratory. Subsequently, the most common dilution value across all three laboratories was determined. For example, 10/198 serum dilution against the HB3 coated plate for all three laboratories was 1 in 25,000.

#### Step 2

The 10/198 dilution determined in Step 1a and Step 1b was used in Step 2 of cELISA. The 10/198 serum dilution obtained from both Step 1a and Step 1b, which had a 2-fold higher concentration was incubated with the competitors (HB3, 3D7, or DiCo3) in plates coated with homologous or heterologous antigens (HB3, 3D7, and DiCo3). In accordance with the materials and methods section, the OD_450nm_ values obtained from the cELISA assay were converted into arbitrary units, and the residual binding of the 10/198 serum was calculated. Figure 3A and 3B illustrate the fraction of 10/198 serum bound against the heterologous or homologous competitor antigens. Figure 3A corresponds to the analysis of Step 2a, where the results were obtained using the 10/198 serum dilution determined based on an OD_450nm_ value of 1.5. On the other hand, Figure 3B represents the analysis of Step 2b, where the same 10/198 serum dilution was used across all three laboratories.

**Figure 3:**
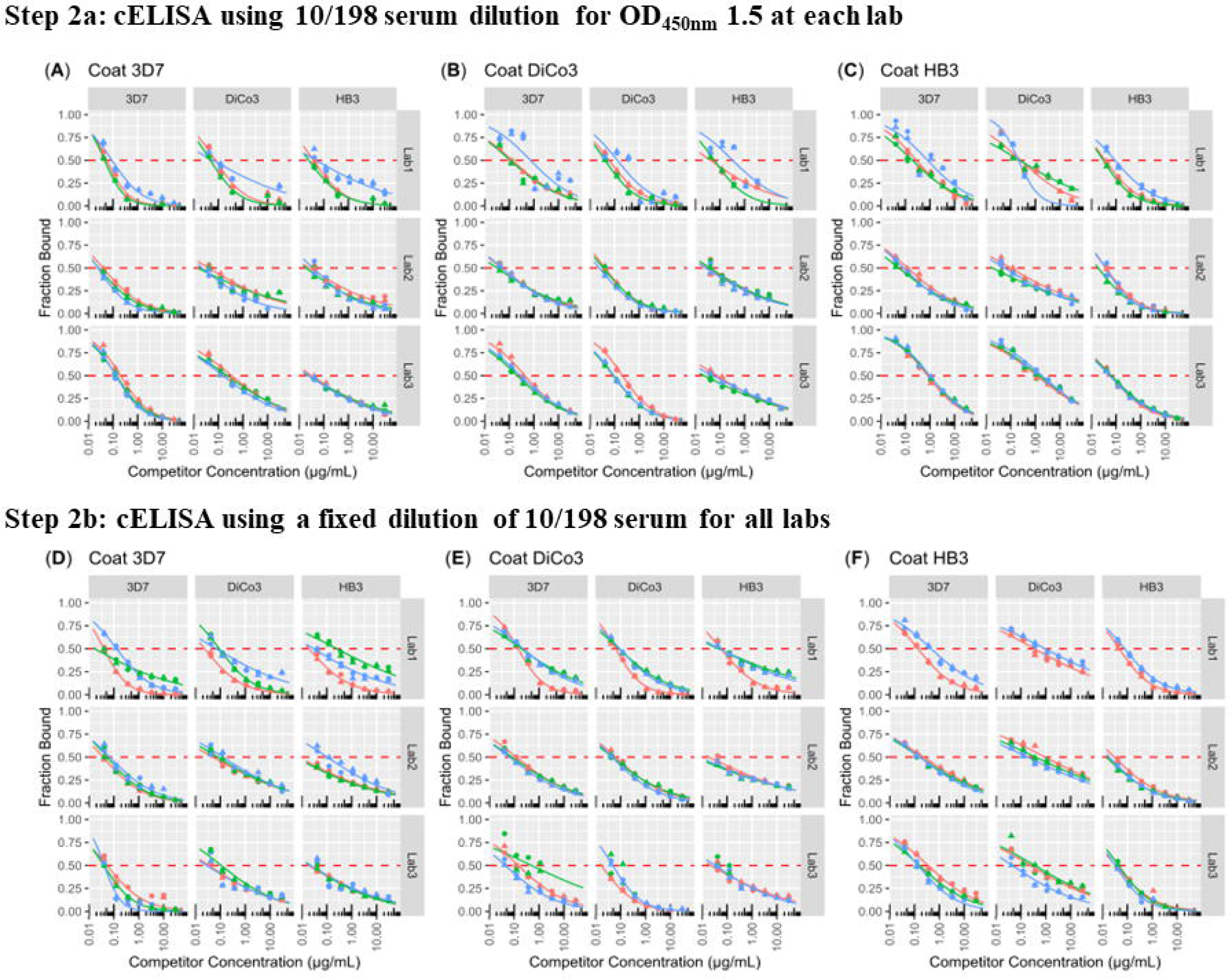
cELISA using 10/198 as serum from stage 1 of the study design: Step 2a (the top graphs); three laboratories performed three independent experiments (n=3, replicates). The 10/198 serum dilution from Step 1 was determined based on the OD_450nm_ 1.5 measurement specific to each laboratory. A 2x dilution of 10/198 was then incubated in an antigen coated well and competed with either homologous or heterologous competitor antigens. Step 2b: The bottom graphs represent the results of three independent experiments (n=3 replicates) in Step 2b. The key difference compared to Step 2a in the cELISA assay is the utilisation of a fixed 10/198 serum dilution. This fixed dilution was determined based on the linear portion of the two-parameter logistics standard curve. The resulting binding interactions were measured and analysed. The IC_50_ value is determined by analysing both, top and bottom graphs and is indicated by the dotted horizontal line on the graphs. The IC_50_ is the concentration of competitor antigen at which half of the binding between the 10/198 serum antibodies and the coated antigen is inhibited. It serves as a measure of the effectiveness of the competitor antigens in competing with the 10/198 antibodies for binding to the coated antigens.

Each laboratory performed three independent experiments, referred to as replicates. However, in the case of Lab 1, as depicted in Figure 3B, one replicate of HB3-coated plates out of the three independent replicates had to be excluded due to technical issues. This exclusion was necessary to ensure the integrity and accuracy of the data analysis and interpretation. Following the competition between the 10/198 serum antibodies and the competitor antigens, the IC_50_ values were calculated to determine the concentration of competitor antigen at which half of the 10/198 serum antibodies remained bound to the coating antigen. These IC_50_ values are depicted by the horizontal dotted lines on the graphs in Fig. 3A and Fig. 3B. The specific IC_50_ values for the antigen-coated plates used in Steps 2a and 2b can be found in Supplementary data 1 and Supplementary Figures 3 and 4.

### Inter-/Intra-lab variations analyses for cELISA Steps 2a and 2b

To understand the inter- and intra-laboratory variations for the IC_50_ values, geometric means for IC_50_ (μg/mL) were calculated. As shown in Figure 4A, the IC_50_ μg/mL obtained from the OD_450nm_ 1.5-based dilution were examined. The results revealed good intra-laboratory agreement especially for the laboratories 2 and 3, whereas laboratory 1 displayed more variations. For example, for DiCo3 coated plates, laboratories 2 and 3 displayed relatively good intra-laboratory agreement, whereas laboratory 1 displays more variations between replicates. When considering HB3-coated plates, laboratories 2 and 3 demonstrated relatively good agreement within the laboratory, whereas laboratory 1 displayed more variation between replicates, with replicate 3 appeared as an outlier. Overall, there is reasonably good intra-laboratory agreement for laboratories 2 and 3 with HB3 competitor, but laboratory 1 displayed more variation between replicates and showed an outlier for both 3D7 and HB3 competitors. The inter-laboratory variation between the laboratories was large, except for the HB3 competitor (Supplementary data 2).

**Figure 4:**
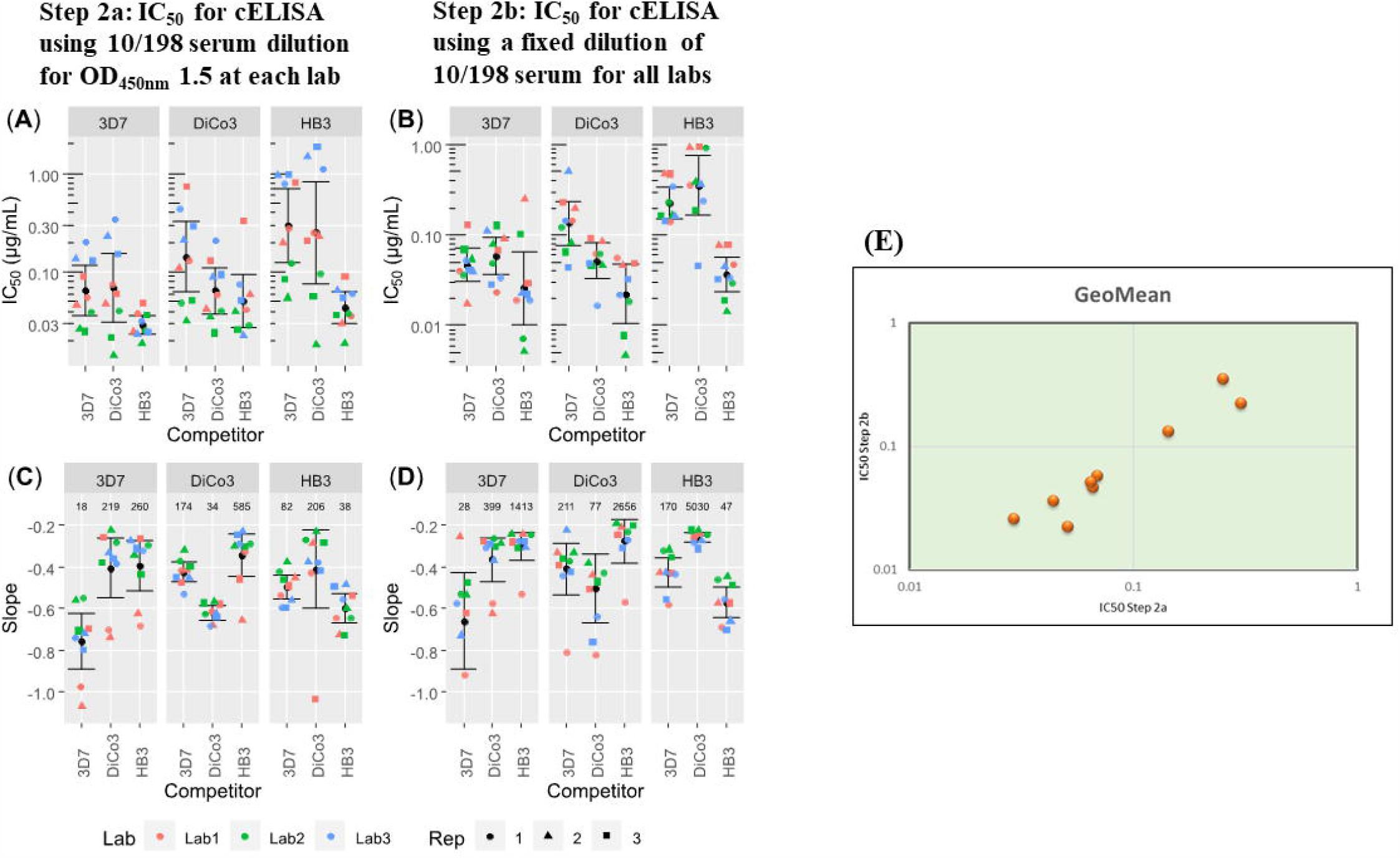
Inter-/Intra-lab variation analyses for cELISA Steps 2a and 2b: All laboratories conducted three independent cELISA experiments for each antigen-coated plate (HB3, 3D7 and DiCo3). From Steps 2a and 2b, the residual binding (μg/mL) of the 10/198 serum was calculated using a two-parameter logistic fit. Subsequently, the IC_50_ (μg/mL) on the y-axis was calculated from the dotted horizontal line as shown in Fig. 3. The inter-and Intra-lab variations and geometric means for IC_50_ (μg/mL) values were calculated to assess the differences between different laboratories and within each laboratory, respectively. (4A), IC_50_ μg/mL for Step 2a; (4B), IC_50_ μg/mL for Step 2b; (C), Slope values for Step 2a and; (4D), Slope values for Step 2b; (E), Correlation of geometric mean between Step 2a and Step 2b.

In Figure 4B, where a fixed dilution of 10/198 was used across all laboratories, the inter- and intra-laboratory variations were further analysed. For the 3D7-coated plates, laboratories 2 and 3 demonstrated relatively close agreement within the laboratory for both 3D7 and DiCo3 competitors. However, laboratory 1 displayed larger variations between replicates for both competitors. When considering the DiCo3-coated plates, laboratories 1 and 2 exhibited relatively close agreement intra-laboratory, while laboratory 3 displayed more variation between replicates for both 3D7 and DiCo3 competitors. Laboratory 1 demonstrated good agreement between replicates, whereas laboratory 2 showed more variation. Laboratory 3, on the other hand, only had two replicates for the HB3 competitor. For the HB3-coated plates, laboratory 2 displayed the best agreement between replicates, while laboratories 1 and 3 showed more variation for the 3D7 competitor. For the DiCo3 competitor, laboratory 1 exhibited the closest agreement between replicates, whereas laboratories 2 and 3 showed more variation between replicates. However, significant inter-laboratory variation remained a notable factor (Supplementary data 2).

Further investigation was conducted to understand the unexpected phenomenon of 10/198 antibodies depleting HB3 more compared to other antigens, even for homologous coat-competitor antigens. To delve deeper into this phenomenon, we examined the IC_50_ μg/mL slopes, as depicted in Figure 4C (10/198 serum dilution based on OD_450nm_ at 1.5) and D (10/198 fixed serum dilution for all laboratories).

As expected, both Figures 4C and D, exhibited steepest slope for homologous coat-competitor. For instance, the 3D7-coated plates when competed with 3D7 antigen (homologous), the slope was steepest for the 3D7 compared to the HB3 or DiCo3 antigens. In contrast, the same homologous coat-competitor (3D7), the IC_50_ μg/mL (Figure 3A), showed the lowest value for HB3 compared to 3D7. Moreover, this pattern of HB3 antigen showing lowest IC_50_ μg/mL persisted for all antigens, irrespective of the coating antigen.

Next, we performed correlation analysis of the geometric mean between Step 2a and 2b to discern whether the IC_50_ μg/mL were different between these two steps (Fig. 4E). The correlation between the step 2a and step 2b was high (r = 0.9585) suggests that either method, whether Step 2a or Step 2b, could be effectively employed to ascertain IC_50_ μg/mL values. All data for the geometric mean calculations and analysis in supplementary data 2.

Based on the results obtained from the stage 1 experiment, where comparable IC_50_ μg/mL were achieved using either the OD_450nm_ 1.5 or a fixed set of dilutions for all laboratories, a fixed set of dilutions was selected for the validation of 10/198 serum in patient samples. This approach ensures consistency and allows for a standardised evaluation of the cELISA results in subsequent stages.

### Use of 10/198 as a standard for plasma samples

For the stage 2 study (Fig. 2), the cELISA validation using patients’ samples was conducted in two laboratories, laboratories 2 and 3, with 10/198 serum serving as the standard. The convalescent patients’ plasma samples, numbered 28 and 32, were tested against three malaria antigens: HB3, 3D7, and DiCo1.

In step 1 of the cELISA, a fixed dilution of the plasma samples was chosen for all three antigens (HB3, 3D7, and DiCo1) across the laboratories. This fixed dilution ensured consistency and comparability of the results obtained.

Moving on to step 2 of the cELISA, each antigen was coated on separate plates and competed with either HB3, 3D7, or DiCo1. The plasma samples, at a 2x concentration, were incubated with the competitor antigens in the plates coated with either homologous or heterologous antigens (HB3, 3D7, and DiCo1). This step allowed for the evaluation of the competition interactions between the patient’s plasma antibodies and the antigens.

As for the standard, 10/198 serum (serially diluted) was incubated without any competitor antigens on the coated antigens. Using the ADAMSEL software and R script, the OD_450nm_ values were converted into arbitrary units, and the residual binding of the plasma samples was calculated. In this study, laboratories 2 and 3 performed two and three independent replicates, respectively. The data analyses for plasma samples 28 and 32 can be found in Supplementary data 3 and Supplementary Figure 5.

In Figure 5, the results for plasma sample 32 showed a high fraction of bound plasma antibodies when competing with heterologous antigens, indicating low competition. This is evident when the plasma sample 32 was coated with antigens HB3, 3D7, and DiCo1 and competed with both homologous and heterologous antigens.

**Figure 5:**
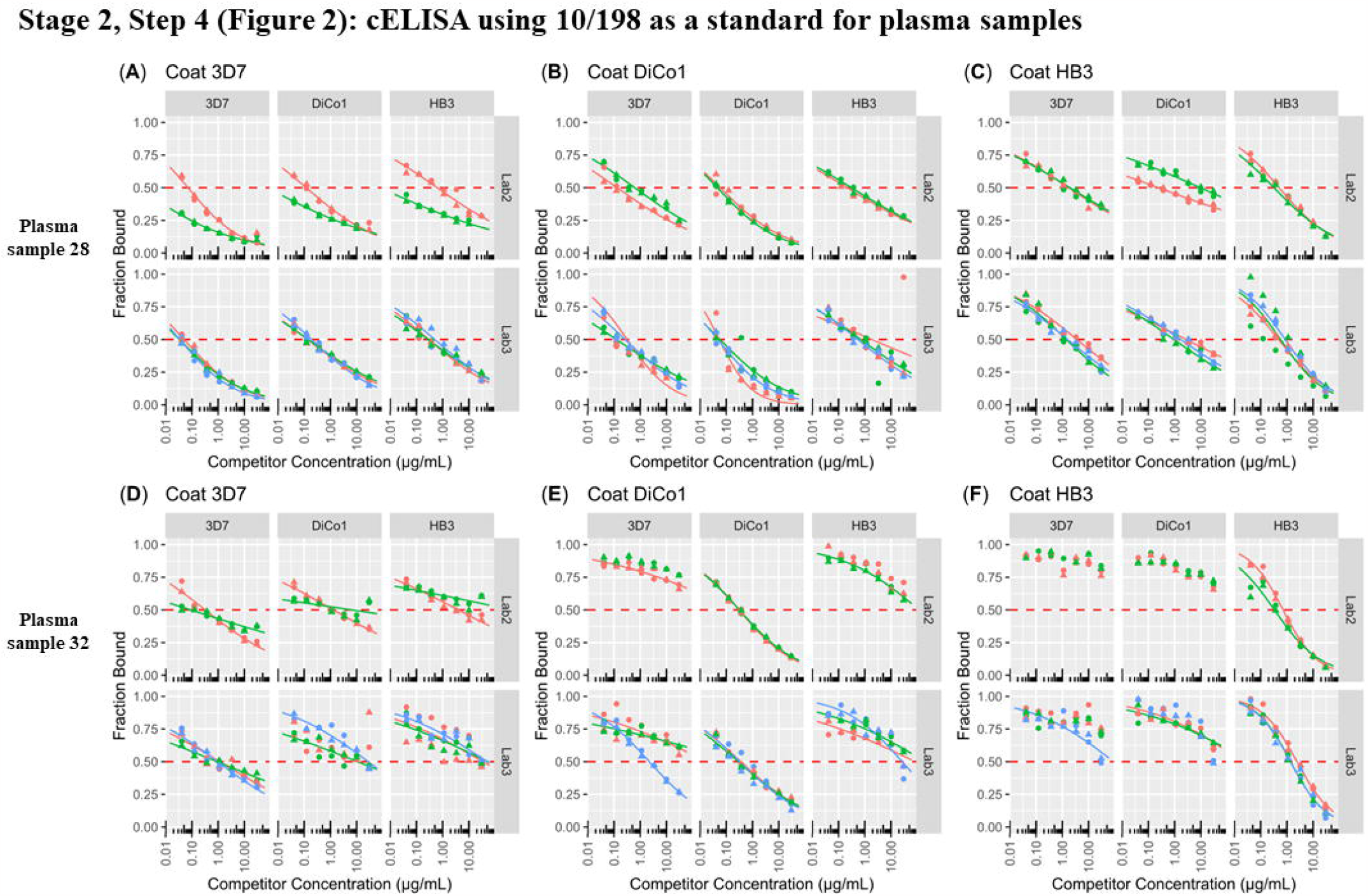
cELISA using 10/198 as a standard and patients’ plasma samples from stage 2 of the study design: Two laboratories, Lab 2 (n=2) and Lab 3 (n=3), performed the cELISA experiments. The plasma samples used in the study were obtained from convalescent patients who had been infected with malaria. The top graphs depict the results obtained using plasma sample 28, while the bottom graphs represent the results obtained using plasma sample 32. These plasma samples were incubated with the coating antigens HB3, 3D7, and DiCo1. The selection of the plasma sample dilution was based on a fixed value determined from the linear portion of the two-parameter logistics standard curve. The 2x diluted plasma samples were then incubated with the competitor antigens in plates coated with either homologous or heterologous antigens. This experimental setup allowed for the assessment of the binding and competition between the plasma antibodies and the different antigens.

### Inter-/Intra-lab variations analyses for plasma samples 32 and 28

To assess the inter- and intra-lab variabilities, IC_50_ values (supplementary data 3 and supplementary figure 5) were calculated based on the graphs in Figure 5. For sample 32, as shown in Figure 6, the IC_50_ values were lowest for homologous antigens, demonstrating the specificity of the cELISA assay. For sample 28, the IC_50_ values were lowest for 3D7-coated plates. Among the three competitor antigens (HB3, 3D7, and DiCo1), both laboratories showed the lowest IC_50_ values for the homologous antigen, followed by DiCo1 (consensus-like AMA1) and the highest for HB3 (South American isolate). Additionally, for DiCo1 coated plates, the lowest IC_50_ values were observed for the homologous antigen in both laboratories, followed by 3D7 and HB3 competitors. HB3-coated plates displayed the lowest IC_50_ values for the homologous antigen, with higher IC_50_ values observed for 3D7 and DiCo1 competitors.

**Figure 6:**
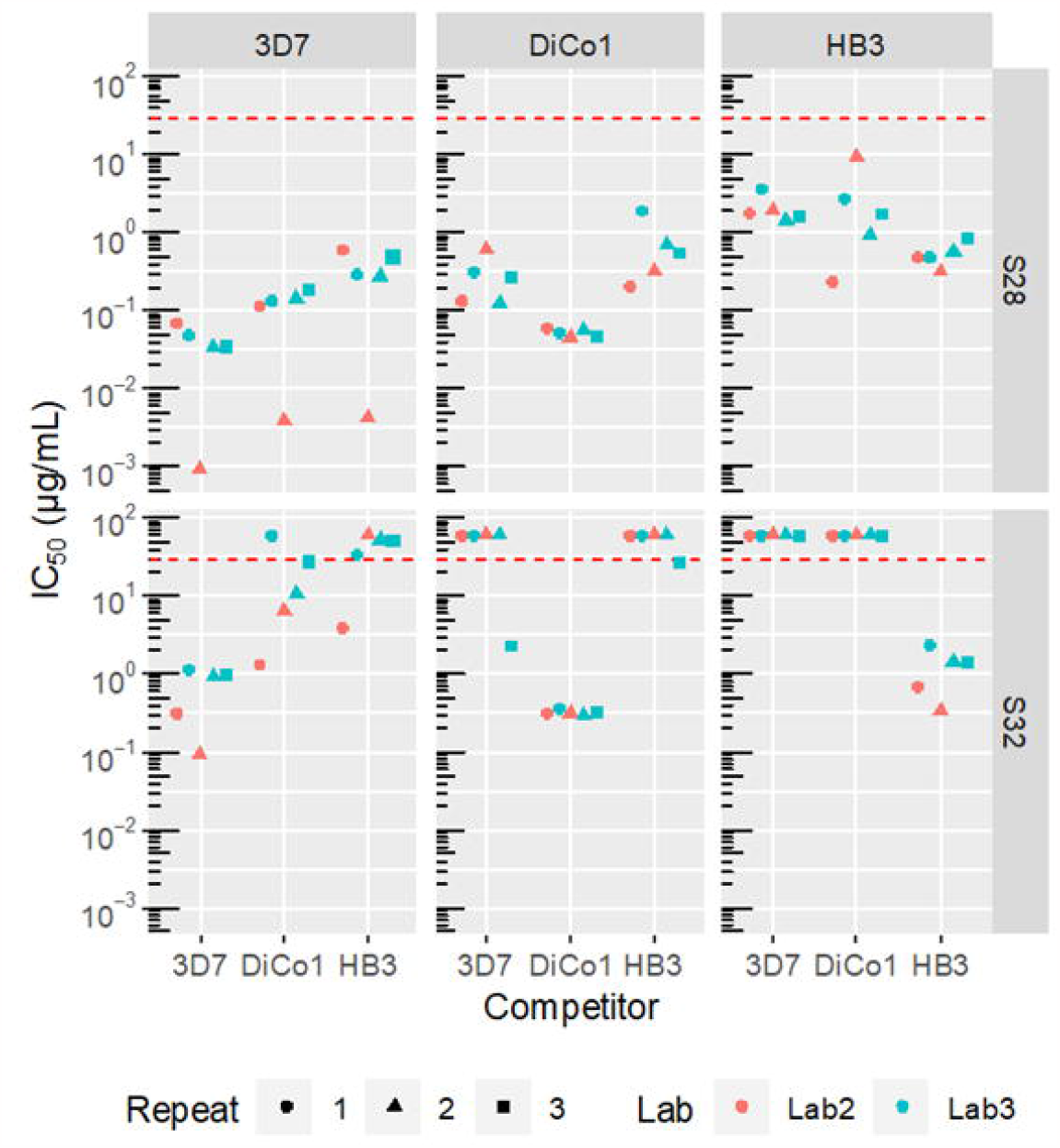
Inter-/Intra-lab variations analyses for plasma samples 32 and 28: The residual binding (μg/mL) of the plasma samples, 28 and 32 were calculated from the two-parameter logistic fit. The IC_50_ (μg/mL) values on the y-axis were then calculated based on the dotted horizontal line as shown in Fig. 5. The x-axis represents competitor antigens used in the experiments, which were incubated with the plasma samples 28 and 32 in wells coated with either homologous or heterologous antigens, HB3, 3D7 and DiCo1. The inter-and Intra-lab variations and geometric means for IC_50_ (μg/ml) were calculated. The experiments were conducted independently by Lab 2 (n=2) and Lab 3 (n=3) for each of the antigen-coated plates (HB3, 3D7, and DiCo1).

These results suggest that the cELISA assay can provide insights into antibody specificity when using samples from a single patient.

## Discussion

The results presented in this paper show that the 10/198 antibody standard is a useful reagent for the harmonisation of antibody assays for AMA1, and its applicability may very well extend beyond AMA1 to other blood stage antigens [17]. It may also be useful for pre-erythrocytic as well as transmission-related antigens, but this requires further investigation.

The results also demonstrate that the cELISA is a difficult to standardise assay as illustrated by relatively large intra- and inter-laboratory reproducibility. Of note here is that intra- and inter-laboratory variation appeared to decrease once operators became more acquainted with the method, which is illustrated by the improved agreement achieved for the plasma samples.

The results presented in this paper show that the 10/198 antibody standard is a useful reagent for the harmonisation of antibody assays for AMA1, and its applicability may very well extend beyond AMA1 to other blood stage antigens [17]. It may also be useful for pre-erythrocytic as well as transmission-related antigens, but this requires further investigation.

The results also demonstrate that the cELISA is a difficult to standardise assay as illustrated by relatively large intra- and inter-laboratory reproducibility. Of note here is that intra- and inter-laboratory variation appeared to decrease once operators became more acquainted with the method, which is illustrated by the improved agreement achieved for the plasma samples. Despite variation within and between laboratories the results reveal similar patterns. The interpretation of the cELISA results for 10/198 as a pooled serum containing antibodies from 149 donors and is expected to cross-react with a wide range of AMA1 variants is not straightforward. The initial expectation is that IC_50_ values for the homologous Coat-Competitor in the cELISA would be the lowest and increase with antigenic distance between the AMA1 variants and slopes would be steepest and increase with antigenic distance between the AMA1 variants. The expectation for the IC_50_ values is lowest for the homologous Coat-Competitor pair, is however, not found. Unexpectedly, the HB3 antigen yielded the lowest IC_50_ values irrespective of coating antigen with the homologous competitor usually as runner up. The slope values, however, agree with the expectations where values are lowest for the homologous Coat-Competitor pair where the increase in competitor concentration required to reduce residual binding from 50 to 10% range between 18 and 77-fold for the homologous pair and between 174 and 5030-fold for heterologous pairs. For the plasma samples the interpretation of the cELISA results is much more straightforward, The IC_50_ is lowest for the homologous coat-competitor pair and slopes for the homologous coat-competitor pair are by far the steepest.

The observed IC_50_ appears less dependent on the OD value than previously reported (Supplementary Figures 1-3) [13]. A possible explanation for this observation may be found in the fact that Kusi et al. [13] used an Alkaline Phosphatase conjugate which is known to be less sensitive (∼10 -fold) than HRP. Therefore, the amount of IgG as detected by a HRP conjugate is likely to be lower than what is detected by an Alkaline Phosphatase conjugate.

The observed IC_50_ appears less dependent on the OD value than previously reported (Supplementary Figures 3-5) [13]. A possible explanation for this observation may be found in the fact that Kusi et al. [13] used an Alkaline Phosphatase conjugate which is known to be less sensitive (∼ 10 -fold) than HRP. Therefore, the amount of IgG as detected by a HRP conjugate is likely to be lower than what is detected by an Alkaline Phosphatase conjugate.

## Conclusions

Although initially developed for interpreting macro data derived from the harmonization of cELISA in this study, ADAMSEL’s applicability extends beyond cELISA and can be leveraged for diverse ELISA types. For this study, the ADAMSEL was used to convert the OD_450nm_ values obtained from the cELISA assay into concentrations expressed as AU using the 10/198 serum as a reference standard.

## Data Availability

All data produced in the present work are contained in the manuscript

## Supplementary materials

Supplementary Figure (contains Figures 1, 2, 3, 4 and 5)

Supplementary data 1, 2 and 3

## Author contributions

Conceptualisation, E.R and P.B.; Methodology, B.K., E.R., P.B and H.McS.; Software, E.R.; Validation, B.K., E.R., P.R., H.S. and D.M.; Formal Analysis, E.R. and B.K.; Investigation, B.K. P.R., H.S. and D.M.; Data Curation, B.K., P.R., H.S., D.M. and E.R.; Writing-Original Draft Preparation, B.K. and E.R.; Writing-Review & Editing, B.K., E.R., P.R., H.S., P.B., and H.McS.; Visualisation, B.K. E.R.; Project Administration, B.K. and E.R.

## Informed Consent Statement

No consent required

