## Supplementary Figures for "Evaluation of 1^st^ WHO Anti-Malaria Reference Reagent for Competition ELISA Harmonisation and Development of ADAMSEL Analytical Platform"

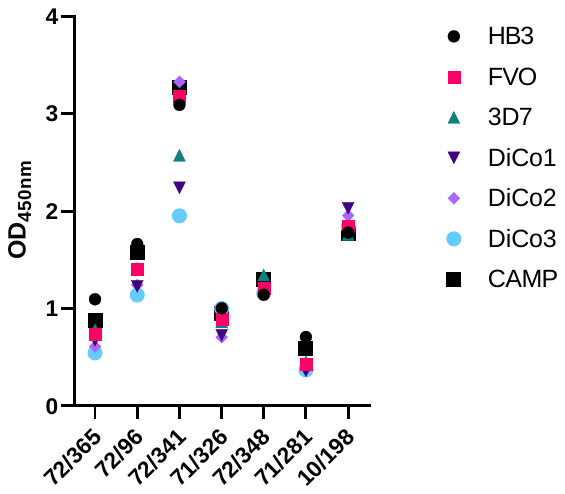

**Supplementary Figure 1: Selection of non-WHO and 1st WHO Reference Reagents for Anti-malaria Assessment.** The non-WHO reference reagents, including product codes 72/365, 72/96, 72/341, 71/326, and 71/281, were obtained from the MHRA's agency website [NIBSC - Products](https://nibsc.org/products.aspx). These reagents were sourced from a single patient with a history of malaria and evaluated for antibodies against specific antigens of P. falciparum and *P. vivax* species. For instance, non-WHO reference reagents with code 71/281 showed positive results for *P. vivax* MSP-119 but negative results for P. falciparum antigens, such as MSP-2 (3D7), AMA1 (3D7), and MSP-119 (K1). Further details can be obtained from the product code 71/281 instructions available on the [NIBSC - Products](https://nibsc.org/products.aspx) website. The 1^st^ WHO reference reagent for antimalaria (*P. falciparum*) human serum, 10/198, was obtained from the pooled plasma of individuals from Kenya and has been detailed in the methods and materials section of the manuscript. Both the non-WHO and 1st WHO reference reagents were screened for antibodies against seven *Pf*AMA1 antigens, including HB3, 3D7, FVO, DiCO1-3, and CAMP. Based on the ELISA OD_450nm_ values, although both 10/198 and 72/341 exhibited high values for all seven antigens, the 10/198 reference reagent was chosen as it represented a pooled serum from Kenyan individuals with a history of malaria.

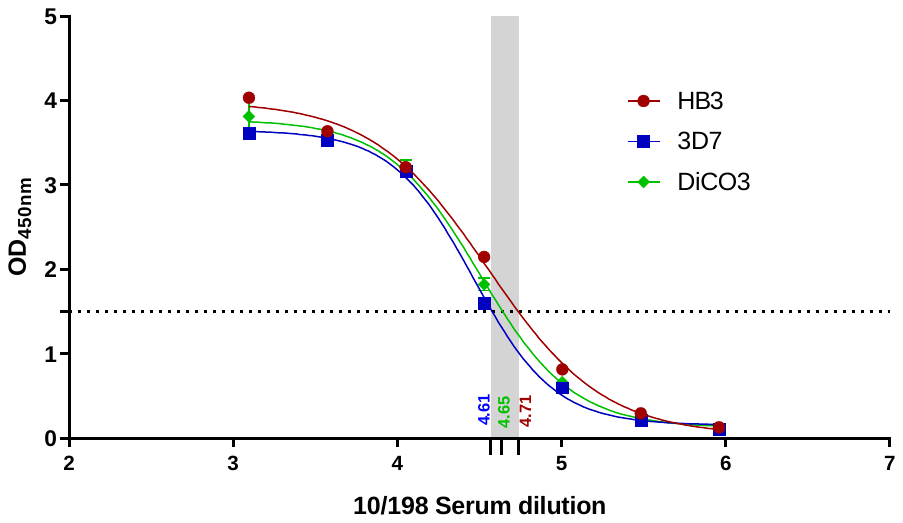

**Supplementary Figure 2: Representative Graph for Serum Dilution in Step 1a (Lab 3).** The 96-well plates were coated with HB3, 3D7 or DiCo3 Ags at a concentration of 2 µg/mL. The 10/198 serum was titrated three-fold, starting from a 1:1250 dilution and then incubated for 2 hours. Subsequently, the plates were washed and added a secondary detection. The development of the plates was carried out using a TMB-blotting solution, and the absorbance was measured at OD_450nm_. The graph represents a four-parameter logistic curve, where the logarithmic serum dilution was plotted on the x-axis, and the corresponding absorbance value at OD_450nm_ was plotted on the y-axis. A dotted horizontal line was drawn at the OD_450nm_ value of 1.5, which was selected as the criterion for the 10/198 serum dilution. As an example, for plates coated with HB3, the log_10_ serum dilution was determined to be 4.71, which corresponds to an exponential value of 1 in 51000 dilutions.

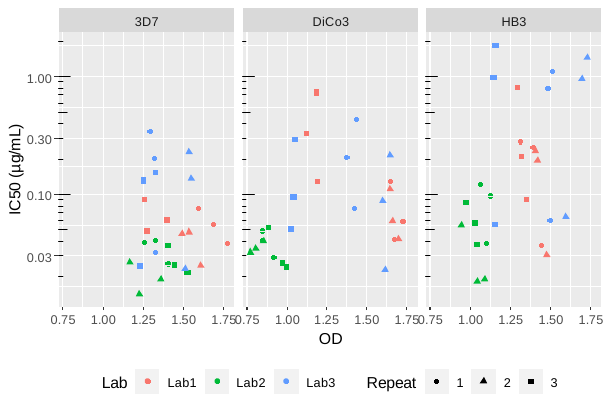

**Supplementary Figure 3: 10/198 IC_50_ versus OD for step 2a**

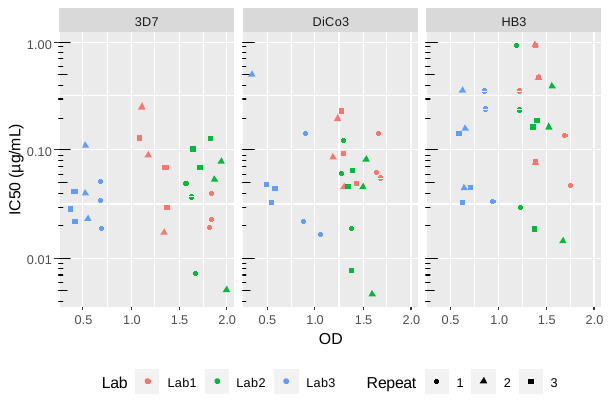

**Supplementary Figure 4.** **10/198 IC_50_ versus OD for step 2b**

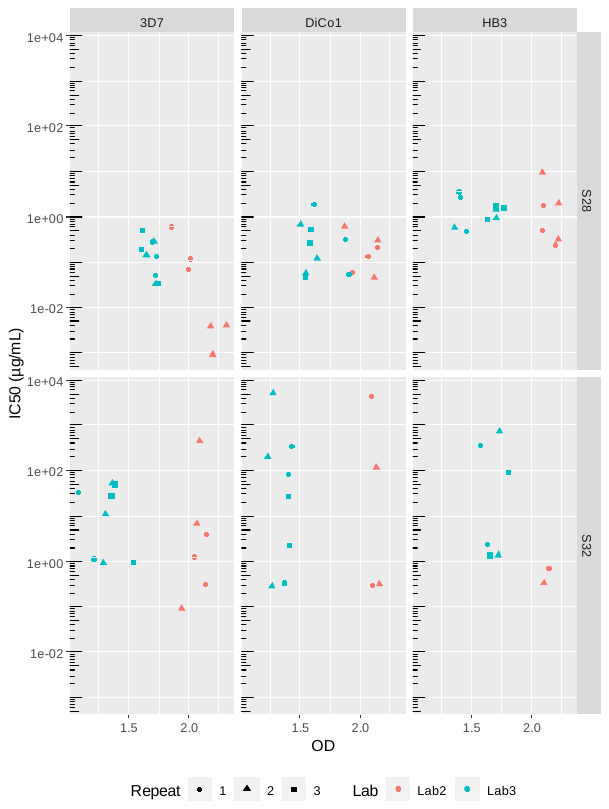

**Supplementary Figure 5. IC_50_ versus OD for plasma samples**
